# Acquisition and Clearance Dynamics of *Campylobacter* in Children in Low- and Middle-Income Countries

**DOI:** 10.1101/2023.02.06.23285359

**Authors:** Dehao Chen, Arie H. Havelaar, James A. Platts-Mills, Yang Yang

## Abstract

**Summary:** 

**Background:** The burden of *Campylobacter* infection is high in children under five years of age in low- and middle-income countries (LMIC), but its acquisition and clearance process is understudied due to scarcity of longitudinal data. We aim to quantify this process using a statistical modeling approach, leveraging data from a multi-nation study.

**Methods:** Motivated by the MAL-ED study in which children from eight low- and middle-income countries were followed up for enteric infections during their first two years of life, we developed a two-stage Markov model to compare the dynamics of acquisition and clearance of *Campylobacter* in children across countries and to explore antibiotic effectiveness on *Campylobacter* clearance. This model was validated using simulations and applied to the longitudinal data from MAL-ED.

**Findings:** The clearance rate was higher than the acquisition rate at most sites and times, but the temporal trend of these rates varied across countries. For *Campylobacter jejuni/coli*, clearance was faster than acquisition under two years of age at all sites. For *Campylobacter* spp., the acquisition rate surpassed the clearance rate in the second half of the first year in Bangladesh, Pakistan and Tanzania, leading to high prevalence in these countries. Bangladesh had the shortest (28 and 57 days) while Brazil had the longest (328 and 306 days) mean times to acquisition for *Campylobacter* spp. and *C. jejuni/coli*, respectively. South Africa had the shortest (10 and 8 days) while Tanzania had the longest (53 and 41 days) mean times to clearance for *Campylobacter* spp. and *C. jejuni/col* respectively. The use of macrolides was associated with accelerated clearance of *C. jejuni/coli* in Bangladesh and Peru and of *Campylobacter* spp. in Bangladesh and Pakistan. The use of fluoroquinolones showed statistically meaningful effectiveness only in Bangladesh but for both *C. jejuni/coli* and *Campylobacter* spp.

**Interpretation:** Higher burden of *Campylobacter* infection was mainly driven by high acquisition rate that was close to or surpassing the clearance rate. Acquisition usually peaked in 11-17 months in the LMIC setting, indicating the importance of targeting the first year of life for effective intervention.

**Funding:** Bill & Melinda Gates Foundation.

## Introduction

Colonization by enteric pathogens, including *Campylobacter*, is a well-documented risk factor for malnutrition in children under five years of age (CU5) in low- and middle-income countries (LMIC) (1,2). The Global Enteric Multicenter Study (GEMS), a case-control study conducted in seven countries in Africa and Asia, found that *Campylobacter* was a leading pathogen responsible for diarrhea in CU5 (3,4). The Etiology, Risk Factors, and Interactions of Enteric Infections and Malnutrition and the Consequences for Child Health and Development (MAL-ED) project was a multi-nation birth cohort study designed to evaluate the association of enteric pathogen infections with malnutrition and further health effects among children under two years of age in eight LMIC countries (5). This study has associated multiple household risk factors with *Campylobacter* infection (6). In addition, along with other recent single-site studies, MAL-ED associated (cumulative) infection of *Campylobacter* with impaired gut functions like gut inflammation and increased intestinal permeability (6–8) and with undernutrition outcomes such as stunted growth and reduced weight gain (2,6,9).

Prevalence of *Campylobacter* usually increases in the early stage of life as infants transit to non-breastmilk food and more activities (6), but the exact temporal patterns of the rate of acquisition (or equivalently, force of infection [FOI]) and the rate of clearance remain under-investigated, mainly due to the sparsity of appropriately designed longitudinal studies. Over the past two decades, only two studies assessed the FOI of *Campylobacter* in CU5 in LMIC’s settings, and the FOI in both studies was estimated by seroconversion rate at the annual scale based on IgG data (10,11). The clearance and acquisition process of *Campylobacter* in CU5 have only been descriptively illustrated in a study in Zanzibar based on a follow-up period of two weeks (12). Less is known about risk/protective factors for acquisition and clearance in LMIC, e.g., the effectiveness of antibiotics on clearance. To date, MAL-ED remains to be the sole completed multisite study that garnered prospective longitudinal data on *Campylobacter* infection and antibiotic use in young children, offering a unique opportunity to quantify such dynamics (2). Here, we present a modeling analysis of the temporal acquisition and clearance dynamics of *Campylobacter* in the MAL-ED study using a simulation-validated two-stage discrete-time Markov process model. We also explored the effectiveness of antibiotics on *Campylobacter* clearance.

## Methods

### Study design, field procedures, and populations

The study design of MAL-ED (enrollment scheme, stool sample collection, laboratory tests and surveillance of antibiotic use) has been presented elsewhere (6,13–15). Briefly, between 2009 and 2014, routine non-diarrheal stool samples were collected monthly during 24 months of the follow-up, in the following eight sites: Haydom, Tanzania; Venda, South Africa; Naushero Feroze, Pakistan; Dhaka, Bangladesh; Vellore, India; Bhaktapur, Nepal; Fortaleza, Brazil; and Loreto, Peru (2,16). During twice-weekly household visits, the reported history of antibiotic use was collected and validated using the packaging of the antibiotics or prescriptions from healthcare providers, and if a caregiver reported a diarrheal episode in a child, the child’s diarrheal sample was garnered for testing (6,15). Diarrheal stool samples might substitute for routine non-diarrheal stool samples if the twice-weekly visit overlapped with a monthly visit. All samples during the 24-month follow-up were assayed by qPCR, which was more sensitive to *Campylobacter jejuni/coli* (16,17). Samples from months 1-12, 15, 18, 21, and 24 were also assayed by enzyme immunosorbent assay (EIA), which was sensitive to a broad group of *Campylobacter* spp. (consisting of *Campylobacter troglodytis, Campylobacter infans, Campylobacter concisus, Campylobacter upsaliensis, C. jejuni*, and *C. coli*) (17,18). Hereafter, we refer to the qPCR- and EIA-detected *Campylobacter* groups as *Campylobacter* spp. and *C. jejuni/coli*, respectively.

### Statistical analysis

We consider a two-stage Markov model with transitions from the state of non-colonized to colonized by *Campylobacter* (acquisition) and vice versa (clearance) among each child. Let *X*_*it*_ indicate the colonization status (1=colonized, 0=not colonized) on day t during the follow-up of the *i*^*th*^ child, where *t* = 0 and *t* = *Ti* correspond to the first and the final follow-up days of the child. We assumed this was a time-inhomogeneous process with the following single-day transition probability matrix:

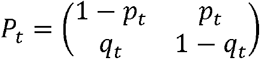

where *p*_*t*_ and *qt* are respectively the daily acquisition probability (0 → 1) and clearance probability (1 → 0) for the transition from day *t* to *t* + 1. Accordingly, 1 − *p*_*t*_ and 1 − *q*_*t*_ are respectively the probabilities of remaining in the non-colonized and colonized states. Hereinafter we use daily acquisition/clearance probability and acquisition/clearance rate interchangeably, as the two are very similar when a daily probability is small. To allow for temporal variation of the transition probabilities, we modeled logit-transformed *p*_*t*_ and *q*_*t*_ as quadratic polynomial functions of time (appendix p1). In the cases where the quadratic term is difficult to identify possibly due to low prevalence, we employed a linear function (appendix p1). For the *C*.

*jejuni/coli* data from Brazil where the observed prevalence was zero in the first two months (Figure 4), we started model fitting from month two to improve identifiability. The likelihood for time-inhomogeneous Markov process is complex. To simplify computation, we approximated the likelihood assuming the transition probabilities were relatively constant between each pair of adjacent observed colonization states. As the time interval between each pair could be as long as three months (6,13,14), we took the average transition probabilities at the two ends and the middle point of each interval (appendix p3). Before applying the model to the MAL-ED data, we validated the identifiability of fundamental parameters in a simulation study (appendix p3). For each of the two *Campylobacter* groups, we also estimated the site-specific and month-dependent mean time to acquisition, mean time to clearance (or mean duration of colonization), and FOI (appendix p1). The FOI during a given period from day *t*_*1*_ to day *t*_*2*_ is calculated as 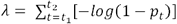, where [*t*1, *t*2] was taken as [0, 365] for year 1, [366, 730] for year 2 and [0, 730] for the two years combined. The FOI is essentially the cumulative hazard. We estimated the effects of recent use of antibiotics up to seven days prior to observed clearance states (appendix p2). The goodness-of-fit of each country-specific model was assessed by comparing the model-simulated trajectory of the prevalence to the observed one (appendix p4).

## Results

The study population sizes with available *Campylobacter* samples were 210, 165, 227, 227, 194, 246, 237, and 209 for the sites in Bangladesh, Brazil, India, Nepal, Peru, Pakistan, South Africa, and Tanzania, respectively. The observed frequencies of the four transition types differed substantially across sites and age years (Figure 1). In the first year of life, colonization activities of *C. jejuni/coli* were represented by the relative frequencies of transitions 0→1, 1→0 and 1→1 were already high in Bangladesh and Tanzania, relatively low in Brazil and South Africa, and moderate in the other four countries (Figure 1A). The colonization activities of *C. jejuni/coli* were increasing from year 1 to year 2 of age in five of the eight countries but the magnitude of increase was more notable again in Bangladesh and Tanzania. The activities were flat in Brazil and dwindled in India and South Africa. In the second year of age, activities in Bangladesh and Tanzania were characterized by excessive sustained colonization indicated by a large proportion of 1→1. As expected, the activity levels of *Campylobacter* spp. in each country and year were higher than those of *Campylobacter jejuni/coli* (Figure 1B). In year 1, Bangladesh, Pakistan and Tanzania had the leading levels of *Campylobacter* spp. colonization activities, followed by India, Nepal, and Peru. These five countries also observed notable increases in the activities from year 1 to year 2, whereas Brazil and South Africa showed no obvious changes over the two years. High levels of sustained colonization of *Campylobacter* spp. in the second year were seen in Bangladesh, Pakistan, and Tanzania.

**Figure 1.**
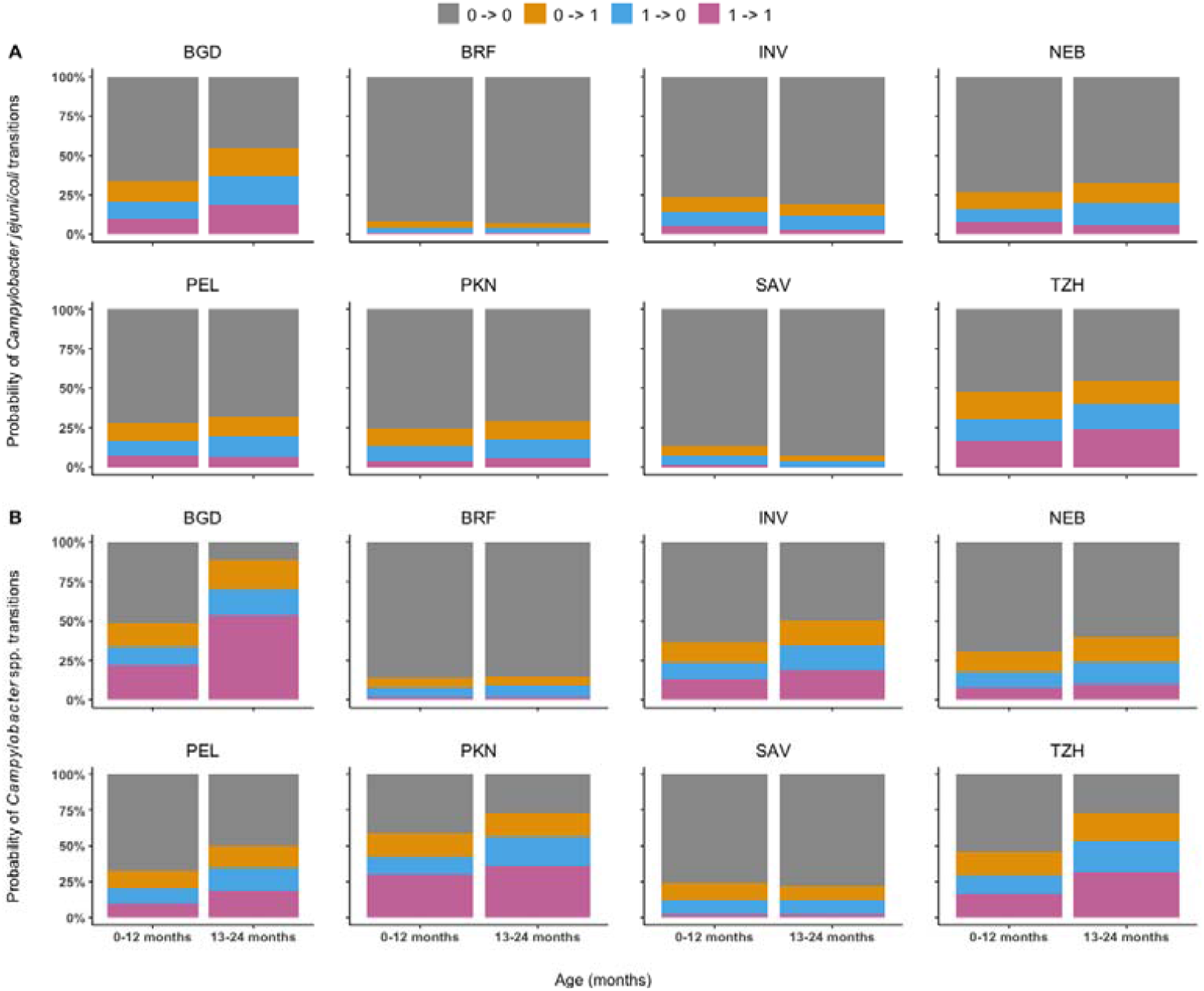
Site-specific relative frequencies of observed transitions of Campylobacter jejuni/coli (A) and Campylobacter spp. (B) between uncolonized and colonized states during 0-12 months (left column) and 13-24 months (right column) of age. Study sites include: Dhaka, Bangladesh (BGD); Vallore, India (INV); Bhaktapur, Nepal (NEB); Naushero Feroze, Pakistan (PKN); Venda, South Africa (SAV); Haydom, Tanzania (TZH); Fortaleza, Brazil (BRF); Loreto, Peru (PEL).

The model-estimated temporal patterns of daily acquisition and clearance probabilities of *C. jejuni/coli* and *Campylobacter* spp. varied considerably across sites with few commonalities (Figure 2). The acquisition probability peaked between months 11 and 17 at most sites for both *Campylobacter* groups with few exceptions. For C. *jejuni/coli*, clearance probabilities were noticeably higher than acquisition probabilities at nearly all sites and all time points except for months 12-16 in Tanzania, when the two probabilities were comparable (Figure 2A). For *Campylobacter* spp., the two probabilities were closer to each other as compared to those for *C. jejuni/coli*, and crossover of the two probabilities occurred (Figure 2B). In Bangladesh, the acquisition probability exceeded the clearance probability around month nine and kept increasing afterwards. In Pakistan and Tanzania, the acquisition probability surpassed the clearance probability slightly towards the end of year 1 but stayed close during year 2. The two opposite transition probabilities for *Campylobacter* spp. were also close during year 2 in India and Peru. In particular, the increase in the clearance probability was steady and relatively steep during the whole two years in South Africa for both *C. jejuni/coli* and *Campylobacter* spp. These model-based results are in line with the summary data in Figure 1 and seem to imply that a high level of colonization activities is associated with two conditions: (1) the acquisition probabilities are relatively high, and (2) the clearance probabilities are lower than or close to the acquisition probabilities. Examples are Bangladesh, Pakistan, India, and Tanzania in Figure 1B and Figure 2B. The estimates of the acquisition and clearance probabilities were derived from the estimates of coefficients in the regression of logit-transformed p_t_ and q_t_ on polynomial terms of time, which are summarized in appendix pp 5-6.

**Figure 2.**
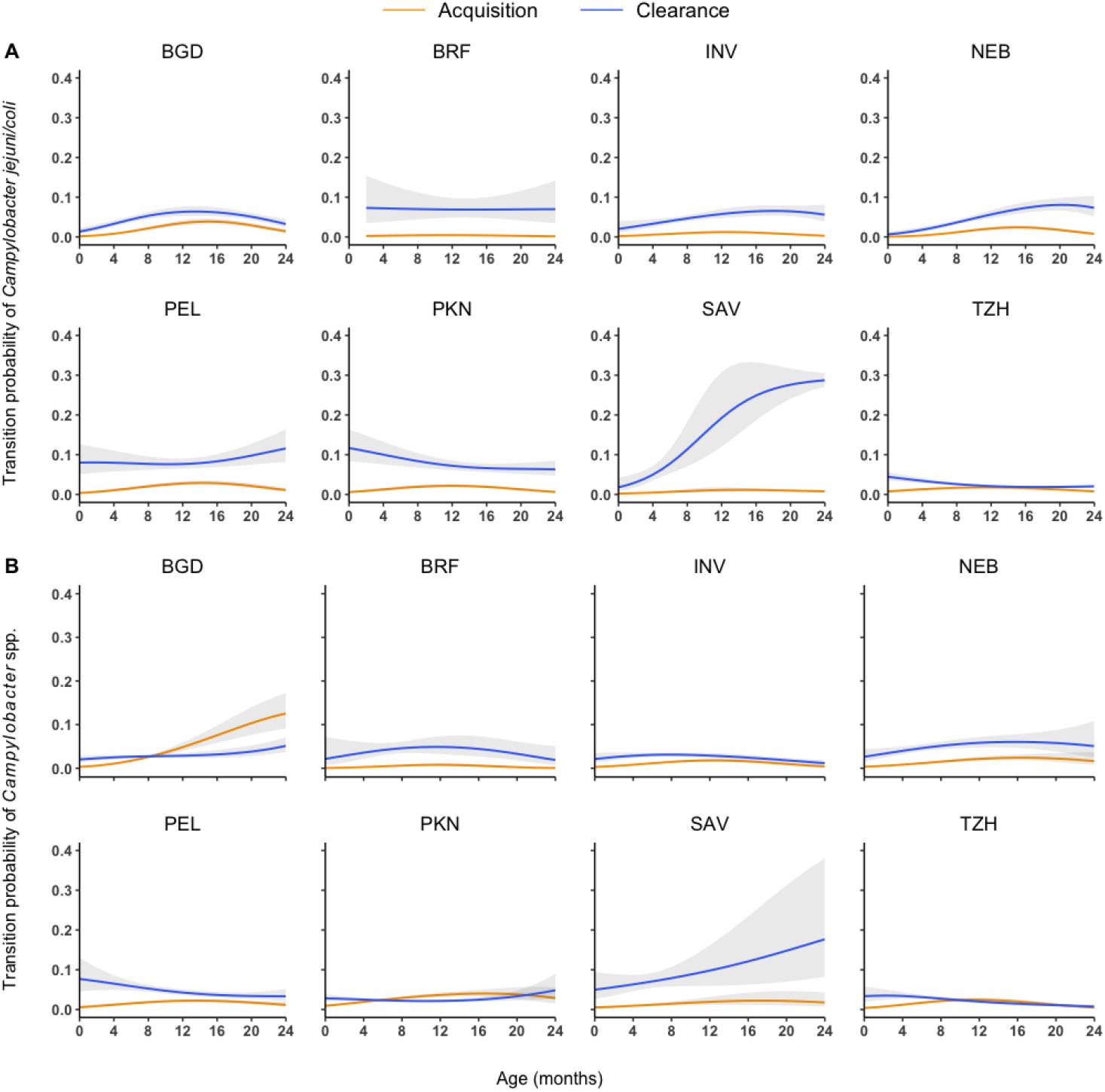
Model-estimated site-specific daily acquisition (orange) and clearance (blue) probabilities of Campylobacter jejuni/coli (A) and Campylobacter spp. (B). 95% asymptotic confidence intervals are shown as gray error bands. Study sites include: Dhaka, Bangladesh (BGD); Vallore, India (INV); Bhaktapur, Nepal (NEB); Naushero Feroze, Pakistan (PKN); Venda, South Africa (SAV); Haydom, Tanzania (TZH); Fortaleza, Brazil (BRF); Loreto, Peru (PEL).

We obtained the mean times to acquisition and to clearance as the reciprocals of the daily acquisition and clearance probabilities respectively (appendix p9). As the probabilities are age-dependent, these duration estimates are also age-dependent. In most countries and for both *Campylobacter* groups, the time to acquisition decreased during year 1 but increased during year 2 with the lowest value near the end of year 1. There is no consistent temporal pattern for the time to clearance. To aid interpretation and comparison, we summarized the real-time daily estimates by taking their median and average over the months (appendix p7). Bangladesh and Brazil respectively had the shortest (28 and 57 days) and the longest (328 and 306 days) mean times to acquisition for both *Campylobacter* spp. and *C. jejuni/coli*; and accordingly, South Africa and Tanzania had the shortest (10 and 8 days) and the longest (53 and 41 days) mean times to clearance. The mean time to clearance of *C. jejuni/coli* was shorter than that of *Campylobacter* spp. in all countries except for Nepal (appendix p7).

Brazil had the lowest whereas Bangladesh had the highest two-year forces of infection (FOI) for both *Campylobacter* groups (appendix p10). For both *Campylobacter* groups, the FOIs of the second year were larger than those of the first year at most sites. As expected, *Campylobacter* spp. had a larger two-year FOI than *C. jejuni/coli* in all countries except for Peru (appendix p10). A reversed order in FOI between *C. jejuni/coli* and *Campylobacter* spp. is possible if *C. jejuni/coli* are the dominant species among *Campylobacter* spp., as then the presumably higher sensitivity of qPCR in comparison to EIA would lead to a higher detection frequency.

MAL-ED surveyed the usages of six antibiotic classes (penicillins, sulfonamides, macrolides, metronidazole, cephalosporins, and fluoroquinolones) (15). We excluded the classes that are known not active against *Campylobacter* from our analyses (19), and macrolides and fluoroquinolones were considered the classes of primary interest.

The use of macrolides or fluoroquinolones during the seven days before the sampling event was significantly associated with increased clearance probability for both *C. jejuni/coli* and *Campylobacter* spp. in Bangladesh (Figure 3). The acceleration factors for clearance (AFC) of *C*.*jejuni/coli* and *Campylobacter* spp. in Bangladesh were estimated as 1.88 (95% CI: 1.58, 2.24) and 1.43 (95% CI: 1.24, 1.64) for macrolides and 1.36 (95% CI: 1.02, 1.83) and 1.35 (95% CI: 1.07, 1.72) for fluoroquinolones. The use of macrolides was also associated with accelerated clearance of *C. jejuni/coli* in Peru (AFC = 1.60, 95% CI: 1.18, 2.18) and *Campylobacter* spp. in Pakistan (AFC = 1.83, 95% CI: 1.36, 2.46) (Figure 3). At most sites, the effects of macrolides on clearance were larger for *C*.*jejuni/coli* than for *Campylobacter* spp., and both *Campylobacter* groups appeared to be more sensitive to macrolides (Figure 3). As detailed in appendix p2, estimates of some antibiotic effects at some sites were not reported due to lack of identifiability as a result of sparse antibiotic use. We translated statistically significant AFC values to relative benefits (ratio of probabilities), where the baseline daily clearance probability in the absence of antibiotics for this calculation was the average baseline daily clearance probability throughout the two-year study period (appendix p11). For example, macrolides reached relative benefits of 5.08 for *Campylobacter* spp. in Pakistan and 4.19 for *C*.*jejuni/coli* in Bangladesh.

**Figure 3.**
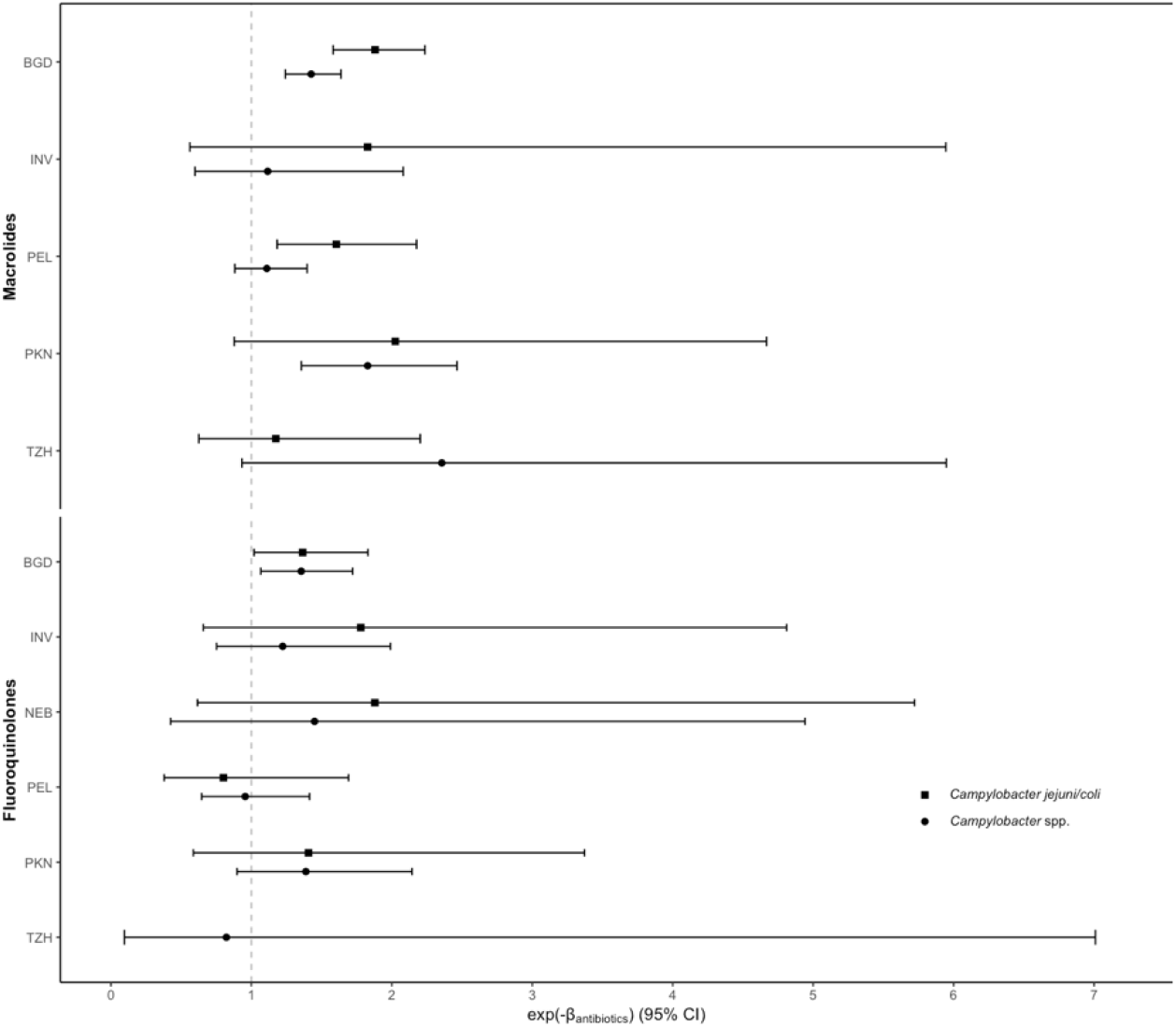
Site-specific effects of macrolides and fluoroquinolones on the clearances of Campylobacter jejuni/coli and Campylobacter spp. Point estimates and 95% confidence intervals of the acceleration factor for clearance (AFC) were shown, which is defined as the ratio of the log probability of clearance with antibiotic use to the log probability of clearance without. Study sites include: Dhaka, Bangladesh (BGD); Vallore, India (INV); Bhaktapur, Nepal (NEB); Naushero Feroze, Pakistan (PKN); Venda, South Africa (SAV); Haydom, Tanzania (TZH); Fortaleza, Brazil (BRF); Loreto, Peru (PEL).

The model-simulated epidemic trajectories of *C. jejuni/coli* and *Campylobacter* spp. were highly consistent with the observed ones at all sites regardless of whether all samples or non-diarrheal samples were modeled, indicating adequate goodness-of-fit (Figure 4). Overall, the prevalence of *C. jejuni/coli* and *Campylobacter* spp. increased sharply in the first year of age, with different peak values and peaking times at different sites, and plateaued or declined gradually during the second year. As clearance of *C. jejuni/coli* was faster than *Campylobacter* spp. in all sites except Nepal (Figure 2, appendix p7), the peak prevalence of *C. jejuni/coli* was generally lower than that of *Campylobacter* spp. (Figure 4). At sites in Bangladesh, Pakistan, and Tanzania where acquisition and clearance rates of *Campylobacter* spp. crossed over (Figure 2), the overall as well as peak prevalence of *Campylobacter* spp. was higher than the other sites (Figure 4). In Bangladesh where acquisition of *Campylobacter* spp. was faster than its clearance in the second year (Figure 2), the peaking of its prevalence was delayed to the end of the second year (Figure 4).

**Figure 4.**
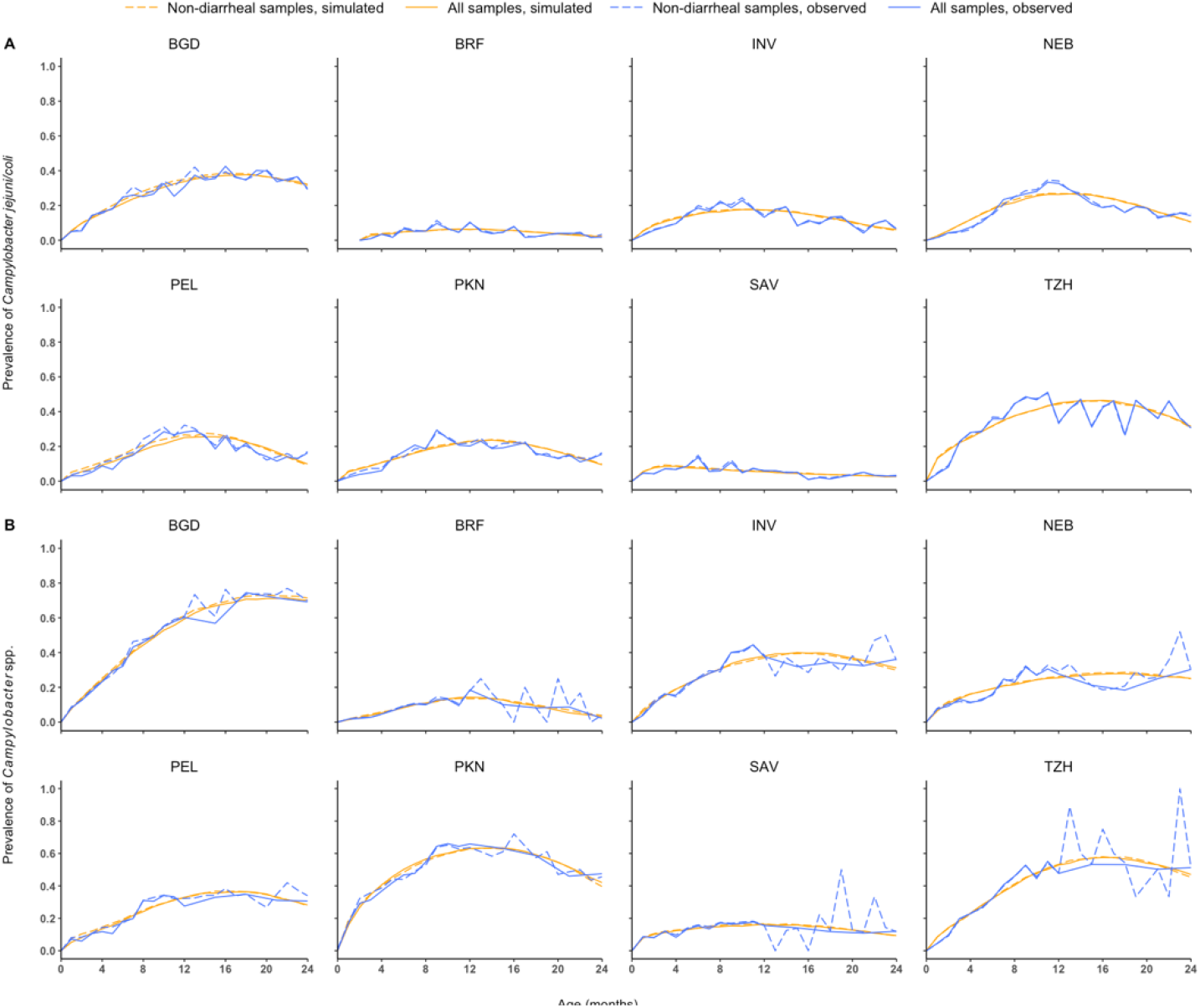
Site-specific observed and simulated longitudinal prevalence trends of Campylobacter jejuni/coli and Campylobacter spp. of all and non-diarrheal samples. The simulated longitudinal prevalence trends were averaged from 100 realizations generated using fundamental parameters estimated from the corresponding observed data.

## Discussion

Harnessing the longitudinal data from MAL-ED, we found that the high burden of *Campylobacter* in young children in LMICs was primarily driven by a relatively high acquisition rate that was close to or exceeding the clearance rate. We also found the acquisition rate was rising during the first year and peaked between months 11 and 17 at most sites for both *C*.*jejuni/coli* and *Campylobacter* spp. in general. The temporal pattern of clearance rate, on the other hand, varied substantially from country to county. The class of macrolides tends to be more active on *C. jejuni/coli* than on *Campylobacter* spp., and both *Campylobacter* groups seem to be more sensitive to macrolides than to fluoroquinolones.

In the first two years of life, while the FOI of *Campylobacter* spp. is often higher than *C. jejuni/coli*, clearance of *C. jejuni/coli* is often faster than *Campylobacter* spp. However, care should be taken when interpretating the comparisons between *C. jejuni/coli* and *Campylobacter* spp., as the former is a subset of the latter. If co-colonization of multiple *Campylobacter* species is common, then our results do not suggest higher FOI or faster clearance of *C. jejuni/coli* compared to other *Campylobacter* species, as it is expected that colonization with any species is faster and clearance of all species is slower when compared to a specific species. On the other hand, if the different species (or species groups such as thermotolerant and non-thermotolerant) compete for ecological niche and do not co-colonize often, then our results can be interpreted as comparisons between *C. jejuni/coli* and other *Campylobacter* species.

Multiple pathogenesis factors (e.g., motility, chemotaxis, adhesion) contribute to the acquisition of *Campylobacter*. (20), triggering innate, humoral, and cell-mediated immune responses in human hosts (21). While recurrent exposures to *Campylobacter* trigger short-term protective effects against symptoms (e.g., diarrhea), it is unclear whether these protective effects accelerate clearance (21). As reflected in our findings, the daily acquisition probability was temporally variant and peaked between 11-17 months at most sites. Future interventions may target the first year when acquisition is accelerating. The non-stopping growth of the acquisition rate of *Campylobacter* spp. throughout the first two years of life in Bangladesh is alarming, likely implying longer efforts needed to reduce *Campylobacter* burden in children. In contrast to acquisition, the trend of the clearance probability of *Campylobacter* appears to be more complicated, increasing with age at some sites but flat or even slightly decreasing at others, possibly related to country differences in maternally acquired immunity in early months, development of human immune system, antibiotics use, and nutrition status (22,23). For example, the steady increase of clearance probability of both *C. jejuni/coli* and *Campylobacter* spp. in South Africa was likely a result of a higher socio-economy level and may explain the less frequent diarrhea and use of antibiotics (15). The different clearance probabilities across countries might be related to different levels of the immunosuppressive effects associated with stunting. For example, stunting prevalence was high in Tanzania and Bangladesh, where clearance probabilities of *Campylobacter* spp. were sometimes lower than acquisition probabilities (24). Slow clearance might also be related to environmental enteric dysfunction, increasing the translocation of pathogen-associated molecular products (PAMPs) such as lipopolysaccharide, weakening host immunity (25).

While the epidemiology of the thermotolerant group *C. jejuni/coli* is well studied given its health impact in the developed world (26), little is known about the non-thermotolerant *Campylobacter* group. Our finding on the stronger antibiotic effects on the clearance of *C. jejuni/coli* than *Campylobacter* spp. may imply the non-thermotolerant group was less sensitive to the two antibiotic classes. Our finding on the larger effect of macrolides on clearance was corroborated by the fact that resistance to fluoroquinolones in *Campylobacter* is more common (27), making macrolides the top empiric medication for campylobacteriosis (28,29), though emerging resistance to macrolides was also documented (30,31).

Our findings are grounded on the novel Markov model validated by a simulation study. Markov models have been applied in studies of acquisition/clearance in a variety of pathogens such as *Neisseria meningitidis* in African countries (32), antibiotic-resistant *Staphylococcus aureus* in a nosocomial setting in the United Kingdom (UK) (33), and *Streptococcus pneumoniae* in UK households (34). Despite the longitudinal nature of these studies, many of them modeled acquisition and clearance as time-invariant, an untenable assumption considering the complex biological interactions among pathogens, hosts and environment. In contrast, our model captures time-varying transition dynamics with a flexible parametric structure while ensuring model identifiability. This framework can be extended to nonparametric structures for more flexibility, e.g., using splines in the regression of the acquisition and clearance probabilities, if supported by abundant data.

A limitation of this modeling study is that the sensitivity and specificity of EIA for *Campylobacter* spp. and qPCR for *C*.*jejuni/coli* were assumed perfect, and misclassification of the colonization status is possible. Another important limitation is that MAL-ED is an observational study, and our findings on the real-world effectiveness of antibiotics on *Campylobacter* clearance should be interpreted with caution. In addition, qPCR targeting *Campylobacter* species other than *C. jejuni/coli* was not performed, and the results with regard to *Campylobacter* spp. cannot be generalized to non-thermotolerant *Campylobacter* species.

In summary, this work uncovered the acquisition and clearance dynamics of *Campylobacter* in children in low-resource settings and the real-world effectiveness of antibiotics on its clearance. Future studies may consider collecting more data on host and environmental characteristics to better explain the heterogeneity in acquisition and clearance patterns across countries. Whenever resources permit, a complete profile of *Campylobacter* at the species-level coupled with multi-species modeling framework would reveal more insights about the acquisition and clearance dynamics of different species and their joint impact on children’s gut health and growth, which will inform future risk assessments and intervention strategies.

## Data Availability

All data produced in the present study are available upon reasonable request to the authors

