## supplementary_appendix for "Acquisition and Clearance Dynamics of *Campylobacter* in Children in Low- and Middle-Income Countries"

### Specification of transition probabilities

To model the temporal variation of the transition probabilities, we link $p_{t}$and$q_{t}$ to quadratic polynomials of time via logit transformations in the following way:

$$u_{t}=logit\left( \frac{p_{t}+q_{t}}{K} \right)=a+bt+ct^{2}$$

$v_{t}=logit\left( \frac{p_{t}}{p_{t}+q_{t}} \right)=d+et+ft^{2}$	(1)

Here $K$ is an upper bound to constrain the sum of $p_{t}$and$q_{t}$ (i.e., $p_{t}+q_{t}<K$) to ensure their values are biologically possible, which is assumed known. As the parameter spaces of $p_{t}$and$q_{t}$ and $\frac{p_{t}}{p_{t}+q_{t}}$ are orthogonal, this parameterization improves identifiability of $p_{t}$and$q_{t}$ and the fundamental parameters $a$, $b$, $c$, $d$, $e,$ and $f$. $p_{t}$ and $q_{t}$ can be expressed in terms of $u_{t}$ and $v_{t}$ via $p_{t}=K*expit\left( u_{t} \right)*expit\left( v_{t} \right)$ and $q_{t}=K*expit\left( u_{t} \right)*\left( 1-expit\left( v_{t} \right) \right)$. We did not model $p_{t}$ and $q_{t}$ directly as polynomials because they are constrained between 0 and 1. A previous SIR-type model estimated the rate of transitioning from full to partial immunity after *Campylobacter* infections to be approximately 13/year, which translates to a daily rate of 13/365$\approx$0·0356 (1). Consequently, the magnitude of $p_{t}$ and $q_{t}$ should be comparable to $1-e^{-0\cdot0356}\approx0\cdot035$. To be conservative, we set $K=$0·3 for the analysis but performed sensitivity analyses with 0.2 and 0.5, and results were not affected (data not shown).

Identifiability issues of $p_{t}$ and $q_{t}$ tended to occur at sites with low overall/peak prevalence, and we troubleshot them in the following ways. In the preliminary analyses of *Campylobacter* spp. at sites in South Africa and Bangladesh, the estimate of $p_{t}+q_{t}$ for the South Africa data could approach $K$with an unstable estimation of the covariance matrix, and the covariance matrix was singular for the Bangladesh data, indicating problems of identifiability. After checking variances of specific parameter estimates and visualizations of $p_{t}$ and $q_{t}$, we chose to simplify $u_{t}$ to a linear function of time, i.e., $u_{t}=a+bt$ for South Africa and Bangladesh. As the site-specific prevalence of *Campylobacter jejuni/coli* was generally lower than *Campylobacter* spp., we observed similar identifiability issues in more sites. To strengthen identifiability for the *Campylobacter jejuni/coli* data, we chose to simplify $u_{t}$ for the data from Brazil, Pakistan, Peru, and Tanzania, and $u_{t}$ and $v_{t}$ for the data from South Africa.

The acquisition interval and duration of colonization are geometric distributions with $p_{t}$ and $q_{t}$ as their corresponding parameters (i.e., acquisition interval = $\frac{1}{p_{t}}$ and duration of colonization = $\frac{1}{q_{t}}$). The overall acquisition intervals and durations of colonization over the two-year follow-up were reported as the medians and geometric means of all $\frac{1}{p_{t}}$ and $\frac{1}{q_{t}}$ of days proximal to a whole month (i.e., $t\in${0, 30, 60, …,720}). Based on the estimated daily $p_{t}$, the force(s) of infection (FOI) during a given period from day $t_{1}$ to day $t_{2}$ is calculated as $\lambda=\sum_{t=t_{1}}^{t_{2}} \left[ -log\left( 1-p_{t} \right) \right]$, which is essentially the cumulative hazard.

### Characterizing effects of recent antibiotic use on clearance

To study the association between recent antibiotic use and the daily clearance probability, we assume the antibiotic use affects $q_{t}$ in the following way:
$q_{t}=\left[ K*expit\left( u_{t} \right)*\left( 1-expit\left( v_{t} \right) \right) \right]^{\exp\left( \beta_{ML}*x_{ML}(t)+\beta_{FQ}*x_{FQ}(t)+\beta_{D}*x_{D}(t) \right)}$	(1)

$x_{ML}(t)$, $x_{FQ}(t)$, and $x_{D}(t)$ were respectively covariates for the use of macrolides (MLs), the use of fluoroquinolones (FQs), and diarrheal status during a n-day window preceding and including day t. Expression (1) is equivalent to a complementary log-log regression, as it can be re-written as

$$\log\left( -\log\left( q_{t} \right) \right)=log\left( -\log\left( K*expit\left( u_{t} \right)*\left( 1-expit\left( v_{t} \right) \right) \right) \right)+\beta_{ML}*x_{ML}\left( t \right)+\beta_{FQ}*x_{FQ}\left( t \right)+\beta_{D}*x_{D}\left( t \right).$$

We chose this parameterization instead of, say, logistic regression, because it seems to be associated with stabler optimization of the likelihood. We considered diarrhea as a confounder, as diarrhea can lead to or be affected by antibiotic treatments (2), and diarrhea is a host response to enteric infection of pathogens. Determination of $x_{ML}(t), x_{FQ}(t), and x_{D}(t)\mathrm{is}$ detailed as follows. If ≥ 1 course(s) of an antibiotic class or ≥ 1 time(s) of diarrhea were initiated during the n-day window prior to and including the day of a sampling event, the covariates = 1; otherwise, the covariates = 0. As antibiotics’ effect is usually short-term after administration, we set $n=8$, assuming the effect last about a week post-use. We conducted a sensitivity analysis for a sixteen-day window and the results were generally similar to those of the eight-day window, with exceptions that additional significant effects of macrolides at the Nepal and Pakistan sites and of fluoroquinolones at the Pakistan site were observed, while the effects of fluoroquinolones at the Bangladesh site were no longer significant under the wider window (Supplementary figure 4).

For both the eight-day and sixteen-day windows, we did not report antibiotic effects at the South Africa and Brazil sites, where the frequencies of using macrolides and fluoroquinolones were too low to make $\beta_{ML}$and$\beta_{FQ}$ identifiable. At the Tanzania site, as the frequencies of fluoroquinolones for *C. jejuni/coli* were too low to make $\beta_{FQ}$ identifiable under both windows, we dropped $x_{FQ}(t)$ from the corresponding models. At the Nepal site under the eight-day window, few macrolides courses were associated with samples positive for *C. jejuni/coli* and *Campylobacter* spp., so we dropped $x_{ML}(t)$ and only evaluated $\beta_{FQ}$ and $\beta_{D}$ in the analysis for the two *Campylobacter* groups. At the Nepal site under the sixteen-day window, given the frequency of using macrolides for *C. jejuni/coli* was too low to yield identifiable $\beta_{ML}$, we only evaluated $\beta_{FQ}$ and $\beta_{D}$ for this scenario.

We present the antibiotic effect in the form of $e^{-\beta}=\frac{\text{log}q_{t}\left( x\left( t \right)=0 \right)}{\text{log}q_{t}\left( x\left( t \right)=1 \right)}$, where $\beta$ can be $\beta_{ML}$ or $\beta_{FQ}$, $x\left( t \right)$ can be $x_{ML}(t)$ or $x_{FQ}(t)$, and$q_{t}\left( x\left( t \right)=1 \right)$ is the value of $q_{t}$ when $x\left( t \right)=1$. An effective antibiotic corresponds to increased clearance probability, i.e., $\frac{q_{t}\left( x\left( t \right)=1 \right)}{q_{t}\left( x\left( t \right)=0 \right)}>1$ and thus $e^{-\beta}>1$. We can view $e^{-\beta}$ as an acceleration factor for the clearance probability, and we name it the acceleration factor for clearance (AFC). For example, when $e^{-\beta}=2$, $q_{t}\left( x\left( t \right)=1 \right)$ is the square-root of $q_{t}\left( x\left( t \right)=0 \right)$. There is no one-to-one mapping between the AFC and other effectiveness measures such as relative risk and odds ratio. The relative risk and odds ratio depend not only on $e^{-\beta}$ but also on the baseline value$q_{t}\left( x\left( t \right)=0 \right)$. To help readers with interpretation, we plot the relative benefit, $\frac{q_{t}\left( x\left( t \right)=1 \right)}{q_{t}\left( x\left( t \right)=0 \right)}$, as a function of $e^{-\beta}$ at different levels of$q_{t}\left( x\left( t \right)=0 \right)$ in Supplementary Figure 3. We call $\frac{q_{t}\left( x\left( t \right)=1 \right)}{q_{t}\left( x\left( t \right)=0 \right)}$ relative benefit instead of relative risk simply because the ratio >1 implies a benefit rather than a risk. When $q_{t}\left( x\left( t \right)=0 \right)=0.2$, $1\leq e^{-\beta}\leq2$ is mapped to $1\leq\frac{q_{t}\left( x\left( t \right)=1 \right)}{q_{t}\left( x\left( t \right)=0 \right)}\leq2$. When $q_{t}\left( x\left( t \right)=0 \right)=0.04$, $1\leq e^{-\beta}\leq2$ is mapped to $1\leq\frac{q_{t}\left( x\left( t \right)=1 \right)}{q_{t}\left( x\left( t \right)=0 \right)}\leq5$.

### Maximum-likelihood inference

Despite the model being described in the form of a single-day transition, the time intervals of most consecutive pairs of the observed colonization events were more than one day given the sampling schedule of MAL-ED (3–5). As $p_{t}$and$q_{t}$ varied from day to day, to simplify computation, we introduced average acquisition and clearance probabilities, respectively denoted by $\bar{p}_{il}$ and $\bar{q}_{il}$, for a multiple-day transition during the $l^{th}$ interval with a duration of $d_{il}$ days for person $i$ such that the transition matrix over the interval can be approximated by:

$$P_{il}=\left( \begin{matrix} 1-\bar{p}_{il} & \bar{p}_{il} \\ \bar{q}_{il} & 1-\bar{q}_{il} \end{matrix} \right)^{d_{il}}$$

We use approximation $\bar{p}_{il}\approx$($p_{t_{i(l-1)}}+p_{t_{i(l-0.5)}}+p_{t_{il}}$)/3 and $\bar{q}_{il}\approx\text{(}q_{t_{i\left( l-1 \right)}}+q_{t_{i(l-0.5)}}+q_{t_{il}}\text{)/3, }$where$\text{ }t_{i(l-1)}$, $t_{i(l-0.5)}$ and $t_{il}$ are the days at the beginning, median, and end of interval $l$ of person $i$. For interval $l$ofperson$i$, let $p_{il}(j,k)$ be the element of $P_{il}$ on row $j$ and column $k$ and $s_{i(l-1)}$ and $s_{il}$ be the infection states at the beginning and end, under the assumption that the colonization processes among children are independent, the likelihood of all data is:

$$L\left( \bar{p}_{il},\bar{q}_{il};y \right)=\prod_{i=1}^{N} \prod_{l=1}^{m_{i}} p_{il}(s_{i(l-1)},s_{il})$$

Where $m_{i}$ denotes the total number of multiple-day transitions of person $i: l\in\left( 1,..,m_{i} \right); N$ denotes the total number of children in a site: $i\in\left( 1,..,N \right)$. We assumed perfect accuracy for the test assays (EIA and PCR) of the two *Campylobacter* groups.

We used maximum likelihood estimation (MLE) for statistical inference. We applied the *BFGS* algorithm to search the optimal log-likelihood for the parameters through the “*optim*” function in R (version 4.1.0), which was also used to perform other analyses. Based on the covariance matrix of the fundamental parameters, we applied the delta method to calculate the asymptotic 95% confidence intervals (CIs) of $p_{t}$ and $q_{t}$. The 95% CIs of $\frac{1}{p_{t}}$ and $\frac{1}{q_{t}}$ and FOI were calculated likewise. For both the MLE and the following model evaluation, we assumed all observations were absent from infection and diarrhea at birth (i.e., zero day of age).

To verify identifiability of the model parameters, we simulated 100 datasets of *Campylobacter* spp. at selected sites (Venda in South Africa and Haydom in Tanzania with low and high prevalence, respectively) using parameters estimated from real data and estimated parameters for each simulated dataset. We examined both bias and mean square error (MSE) for the coefficients ($a, b, c, d, e, f$) in expression (1). Overall, parameters were identified accurately with small bias and variance when data are abundant, e.g., when the true parameters were based on the Tanzania data where prevalence was high (Supplementary table 4, p8). When using parameters estimated from the South African data where prevalence was low, only one parameter ($a$) showed moderate bias, and all parameters had larger MSE than the Tanzania scenario. The median of the estimates for $a$ is however close to the true value. Overall, the proposed model is identifiable with adequate precision.

### Model evaluation

Based on the estimated fundamental parameters ($a,b,c,d,e,f$) and their covariance matrix specific to each site and each *Campylobacter* group, we resampled 100 sets of the fundamental parameters and simulated epidemics for each setting. The epidemics were simulated by sampling the colonization state on the later day of each single-day transition using a multinomial distribution conditioning on the colonization state on the corresponding earlier day of each transition. The simulated prevalence at the end of each month over the 24-month follow-up period was saved to represent the longitudinal trajectory. Goodness-of-fit of the model was assessed by comparing the mean curve of the 100 simulated trajectories of prevalence to the observed prevalence values. As the number of diarrheal samples was non-trivial (6), we fitted models and assessed goodness of fit for both all samples (diarrhea-triggered samples and routine surveillance samples) and routine surveillance samples to investigate if diarrhea potentially introduced bias to the estimates.

**Supplementary table 1** Fundamental parameters a, b, c estimated from full and non-diarrheal *Campylobacter* spp. and *Campylobacter* *jejuni/coli* data.

| Sites^*^ | *Campylobacter*  Groups | Samples | Regression Coefficients (95% CI) | | |
| --- | --- | --- | --- | --- | --- |
|  |  |  | $a$ | $b$ | $c$ |
| BGD | *C.* *jejuni/coli* | All | -2.93 (-3.46, -2.41) | 7.62 (5.13, 10.11) | -6.38 (-8.61, -4.15) |
|  |  | Non-diarrheal | -2.59 (-3.11, -2.07) | 3.55 (1.42, 5.67) | -2.90 (-4.77, -1.03) |
|  | *Campylobacter* spp. | All | -2.48 (-2.82, -2.15) | 2.85 (1.83, 3.86) | -^&^ |
|  |  | Non-diarrheal | -2.62 (-3.02, -2.23) | 1.85 (0.61, 3.09) | -^&^ |
| BRF | *C. jejuni/coli* | All | -1.09 (-2.22, 0.05) | -0.08 (-1.93, 1.77) | -^&^ |
|  |  | Non-diarrheal | -1.30 (-2.42, -0.19) | 0.10 (-1.69, 1.90) | -^&^ |
|  | *Campylobacter* spp. | All | -2.53 (-3.81, -1.25) | 4.43 (-1.02, 9.89) | -4.57 (-9.22, 0.08) |
|  |  | Non-diarrheal | -2.20 (-3.58, -0.81) | 2.95 (-2.94, 8.84) | -3.71 (-8.80, 1.37) |
| INV | *C.* *jejuni/coli* | All | -2.52 (-3.20, -1.83) | 4.18 (1.25, 7.12) | -3.07 (-5.63, -0.51) |
|  |  | Non-diarrheal | -2.34 (-3.06, -1.62) | 2.74 (-0.23, 5.72) | -1.96 (-4.54, 0.61) |
|  | *Campylobacter* spp. | All | -2.44 (-2.95, -1.93) | 3.48 (1.21, 5.75) | -3.94 (-5.97, -1.90) |
|  |  | Non-diarrheal | -2.71 (-3.25, -2.17) | 3.56 (1.22, 5.89) | -4.37 (-6.42, -2.33) |
| NEB | *C.* *jejuni/coli* | All | -3.74 (-4.46, -3.02) | 8.00 (5.06, 10.94) | -5.25 (-7.76, -2.75) |
|  |  | Non-diarrheal | -3.32 (-4.09, -2.56) | 5.79 (2.77, 8.81) | -3.64 (-6.18, -1.11) |
|  | *Campylobacter* spp. | All | -2.21 (-2.75, -1.67) | 3.80 (1.12, 6.48) | -2.83 (-5.69, 0.02) |
|  |  | Non-diarrheal | -2.48 (-3.05, -1.92) | 4.79 (2.28, 7.30) | -5.55 (-7.80, -3.30) |
| PEL | *C.* *jejuni/coli* | All | -0.95 (-1.57, -0.33) | 0.64 (-0.48, 1.75) | -^&^ |
|  |  | Non-diarrheal | -1.75 (-2.19, -1.31) | 0.60 (-0.15, 1.35) | -^&^ |
|  | *Campylobacter* spp. | All | -0.97 (-1.69, -0.25) | -0.53 (-3.35, 2.29) | -0.24 (-2.72, 2.24) |
|  |  | Non-diarrheal | -1.84 (-2.47, -1.21) | 0.58 (-2.09, 3.24) | -1.71 (-4.09, 0.66) |
| PKN | *C.* *jejuni/coli* | All | -0.37 (-0.94, 0.20) | -0.83 (-1.67, 0.01) | -^&^ |
|  |  | Non-diarrheal | -0.86 (-1.52, -0.20) | -0.66 (-1.58, 0.26) | -^&^ |
|  | *Campylobacter* spp. | All | -1.95 (-2.28, -1.62) | 1.20 (-0.64, 3.05) | -0.31 (-2.51, 1.89) |
|  |  | Non-diarrheal | -2.41 (-2.79, -2.04) | 2.80 (0.97, 4.62) | -3.67 (-5.39, -1.94) |
| SAV | *C.* *jejuni/coli* | All | -2.63 (-3.52, -1.73) | 6.75 (2.38, 11.13) | -^&^ |
|  |  | Non-diarrheal | -2.38 (-3.46, -1.30) | 5.36 (-0.02, 10.74) | -^&^ |
|  | *Campylobacter* spp. | All | -1.49 (-2.26, -0.73) | 2.11 (-0.73, 4.94) | -^&^ |
|  |  | Non-diarrheal | -1.50 (-2.18, -0.81) | 1.59 (-0.73, 3.91) | -^&^ |
| TZH | *C.* *jejuni/coli* | All | -1.56 (-1.83, -1.29) | -0.71 (-1.13, -0.29) | -^&^ |
|  |  | Non-diarrheal | -1.64 (-1.92, -1.36) | -0.60 (-1.03, -0.17) | -^&^ |
|  | *Campylobacter* spp. | All | -1.95 (-2.55, -1.35) | 2.00 (-0.68, 4.67) | -3.18 (-5.52, -0.84) |
|  |  | Non-diarrheal | -1.95 (-2.57, -1.32) | 1.59 (-1.16, 4.33) | -2.94 (-5.31, -0.56) |

^*^Sites included: Dahka, Bangladesh (BGD); Vallore, India (INV); Bhaktapur, Nepal (NEB); Naushero Feroze, Pakistan (PKN); Venda, South Africa (SAV); Haydom, Tanzania (TZH); Fortaleza, Brazil (BRF); Loreto, Peru (PEL).

^&^Given preliminary analysis indicated an identifiability issue existed when parameter c was kept in the model for this setting, this parameter was excluded in formal analysis.

**Supplementary table 2** Fundamental parameters d, e, f estimated from full and non-diarrheal *Campylobacter* spp. and *Campylobacter* *jejuni/coli* data.

| Sites^*^ | *Campylobacter* groups | Samples | Regression Coefficients (95% CI) | | |
| --- | --- | --- | --- | --- | --- |
|  |  |  | $d$ | $e$ | $f$ |
| BGD | *C.* *jejuni/coli* | All | -2.13 (-2.49, -1.76) | 4.86 (3.47, 6.24) | -3.57 (-4.77, -2.37) |
|  |  | Non-diarrheal | -2.20 (-2.60, -1.79) | 4.87 (3.32, 6.42) | -3.50 (-4.86, -2.14) |
|  | *Campylobacter* spp. | All | -1.89 (-2.22, -1.56) | 6.87 (5.53, 8.20) | -4.08 (-5.28, -2.89) |
|  |  | Non-diarrheal | -1.77 (-2.16, -1.38) | 6.36 (4.71, 8.01) | -3.77 (-5.27, -2.27) |
| BRF | *C. jejuni/coli* | All, | -3.73 (-4.67, -2.79) | 4.20 (0.42, 7.98) | -4.34 (-7.77, -0.92) |
|  |  | Non-diarrheal | -4.00 (-5.04, -2.97) | 5.26 (1.12, 9.41) | -5.42 (-9.19, -1.65) |
|  | *Campylobacter* spp. | All | -3.58 (-4.39, -2.76) | 7.03 (3.52, 10.53) | -7.09 (-10.50, -3.68) |
|  |  | Non-diarrheal | -3.91 (-4.74, -3.07) | 8.78 (5.01, 12.54) | -9.26 (-13.15, -5.36) |
| INV | *C.* *jejuni/coli* | All | -2.24 (-2.69, -1.78) | 3.36 (1.53, 5.18) | -4.02 (-5.68, -2.36) |
|  |  | Non-diarrheal | -2.34 (-2.81, -1.88) | 3.69 (1.79, 5.58) | -4.27 (-6.01, -2.53) |
|  | *Campylobacter* spp. | All | -2.02 (-2.40, -1.64) | 5.19 (3.58, 6.80) | -4.21 (-5.71, -2.71) |
|  |  | Non-diarrheal | -1.97 (-2.41, -1.53) | 5.23 (3.34, 7.12) | -4.37 (-6.15, -2.58) |
| NEB | *C.* *jejuni/coli* | All | -2.26 (-2.83, -1.70) | 5.09 (3.10, 7.08) | -5.05 (-6.68, -3.42) |
|  |  | Non-diarrheal | -2.44 (-3.01, -1.86) | 5.53 (3.48, 7.58) | -5.35 (-7.05, -3.66) |
|  | *Campylobacter* spp. | All | -2.06 (-2.41, -1.71) | 3.28 (1.82, 4.73) | -2.34 (-3.67, -1.02) |
|  |  | Non-diarrheal | -2.03 (-2.43, -1.63) | 3.16 (1.42, 4.91) | -2.36 (-4.02, -0.70) |
| PEL | *C.* *jejuni/coli* | All | -3.09 (-3.42, -2.76) | 7.55 (6.22, 8.88) | -6.81 (-8.01, -5.60) |
|  |  | Non-diarrheal | -3.26 (-3.69, -2.83) | 7.74 (6.07, 9.42) | -6.83 (-8.33, -5.34) |
|  | *Campylobacter* spp. | All | -2.59 (-2.89, -2.29) | 6.14 (4.84, 7.44) | -4.59 (-5.81, -3.37) |
|  |  | Non-diarrheal | -2.69 (-3.10, -2.27) | 6.64 (4.82, 8.47) | -5.17 (-6.92, -3.43) |
| PKN | *C.* *jejuni/coli* | All | -3.00 (-3.28, -2.71) | 6.53 (5.33, 7.72) | -5.84 (-6.97, -4.72) |
|  |  | Non-diarrheal | -2.99 (-3.33, -2.65) | 6.54 (5.11, 7.96) | -5.96 (-7.29, -4.62) |
|  | *Campylobacter* spp. | All | -1.11 (-1.35, -0.87) | 5.98 (4.90, 7.05) | -5.38 (-6.39, -4.37) |
|  |  | Non-diarrheal | -1.03 (-1.34, -0.71) | 6.10 (4.65, 7.55) | -5.81 (-7.26, -4.36) |
| SAV | *C.* *jejuni/coli* | All | -2.10 (-2.41, -1.79) | -1.49 (-2.05, -0.93) | -^&^ |
|  |  | Non-diarrheal | -2.19 (-2.51, -1.87) | -1.45 (-2.03, -0.86) | -^&^ |
|  | *Campylobacter* spp. | All | -2.28 (-2.61, -1.95) | 2.65 (1.12, 4.17) | -2.67 (-4.18, -1.16) |
|  |  | Non-diarrheal | -2.31 (-2.64, -1.97) | 2.59 (1.02, 4.16) | -2.62 (-4.18, -1.07) |
| TZH | *C.* *jejuni/coli* | All | -1.75 (-2.03, -1.47) | 5.52 (4.27, 6.76) | -4.73 (-5.92, -3.54) |
|  |  | Non-diarrheal | -1.75 (-2.04, -1.46) | 5.49 (4.22, 6.76) | -4.69 (-5.89, -3.49) |
|  | *Campylobacter* spp. | All | -2.18 (-2.53, -1.83) | 7.80 (6.24, 9.37) | -6.07 (-7.57, -4.58) |
|  |  | Non-diarrheal | -2.14 (-2.50, -1.78) | 7.66 (6.04, 9.27) | -5.99 (-7.54, -4.44) |

^*^Sites included: Dahka, Bangladesh (BGD); Vallore, India (INV); Bhaktapur, Nepal (NEB); Naushero Feroze, Pakistan (PKN); Venda, South Africa (SAV); Haydom, Tanzania (TZH); Fortaleza, Brazil (BRF); Loreto, Peru (PEL).

^&^Given preliminary analysis indicated an identifiability issue existed when parameter f was kept in the model for this setting, this parameter was excluded in formal analysis.

**Supplementary table 3** Model-estimated mean time to acquisition and mean time to clearance (or duration of colonization) of *Campylobacter* *jejuni/coli* and *Campylobacter* spp. in the first two years of age. These estimates were obtained by taking the median and average of the model-estimated real-time mean times to acquisition or clearance over the study period.

| Sites^*^ |  | Median  (95% CI) | Mean  (95% CI) |
| --- | --- | --- | --- |
|  | Time to acquisition (days) |  |  |
| BGD | *C. jejuni/coli* | 39 (33, 47) | 57 (50, 64) |
|  | *Campylobacter* spp. | 21 (17, 25) | 28 (24, 33) |
| BRF | *C. jejuni/coli* | 274 (177, 422) | 306 (215, 437) |
|  | *Campylobacter* spp. | 252 (137, 462) | 328 (229, 468) |
| INV | *C. jejuni/coli* | 125 (101, 155) | 143 (125, 164) |
|  | *Campylobacter* spp. | 88 (67, 114) | 100 (86, 116) |
| NEB | *C. jejuni/coli* | 70 (59, 83) | 104 (92, 118) |
|  | *Campylobacter* spp. | 54 (44, 66) | 68 (55, 84) |
| PEL | *C. jejuni/coli* | 49 (40, 61) | 60 (51, 71) |
|  | *Campylobacter* spp. | 57 (44, 74) | 64 (55, 75) |
| PKN | *C. jejuni/coli* | 62 (51, 74) | 72 (61, 84) |
|  | *Campylobacter* spp. | 30 (26, 35) | 35 (30, 42) |
| SAV | *C. jejuni/coli* | 108 (83, 140) | 132 (102, 170) |
|  | *Campylobacter* spp. | 55 (35, 86) | 67 (45, 99) |
| TZH | *C. jejuni/coli* | 70 (62, 79) | 75 (68, 83) |
|  | *Campylobacter* spp. | 62 (48, 79) | 74 (64, 86) |
|  | Time to clearance (days) |  |  |
| BGD | *C. jejuni/coli* | 20 (17, 23) | 23 (20, 26) |
|  | *Campylobacter* spp. | 35 (28, 43) | 33 (28, 39) |
| BRF | *C. jejuni/coli* | 14 (8, 25) | 14 (10, 20) |
|  | *Campylobacter* spp. | 25 (15, 44) | 28 (20, 38) |
| INV | *C. jejuni/coli* | 18 (12, 26) | 21 (18, 24) |
|  | *Campylobacter* spp. | 39 (31, 49) | 42 (37, 49) |
| NEB | *C. jejuni/coli* | 18 (15, 21) | 25 (21, 29) |
|  | *Campylobacter* spp. | 18 (10, 32) | 20 (16, 25) |
| PEL | *C. jejuni/coli* | 12 (9, 18) | 12 (10, 14) |
|  | *Campylobacter* spp. | 23 (20, 28) | 22 (19, 25) |
| PKN | *C. jejuni/coli* | 14 (12, 16) | 13 (11, 15) |
|  | *Campylobacter* spp. | 40 (34, 47) | 37 (32, 44) |
| SAV | *C. jejuni/coli* | 5 (3, 8) | 8 (6, 10) |
|  | *Campylobacter* spp. | 10 (6, 17) | 10 (7, 15) |
| TZH | *C. jejuni/coli* | 48 (42, 53) | 41 (37, 46) |
|  | *Campylobacter* spp. | 50 (40, 63) | 53 (45, 61) |

*Sites included: Dahka, Bangladesh (BGD); Vallore, India (INV); Bhaktapur, Nepal (NEB); Naushero Feroze, Pakistan (PKN); Venda, South Africa (SAV); Haydom, Tanzania (TZH); Fortaleza, Brazil (BRF); Loreto, Peru (PEL).

**Supplementary table 4** Model validation: median, inter-quartile range (IQR), mean and mean square error (MSE) of estimated model coefficients ($a$, $b$, $c$, $d$, $e,$ and $f$) over 100 simulated datasets. Data-generating parameters were estimated from the *Campylobacter* spp. data from sites in South Africa and Tanzania.

| Sites^*^ |  | *a* | *b* | *c* | *d* | *e* | *f* |
| --- | --- | --- | --- | --- | --- | --- | --- |
| SAV | True parameters  (95% CI)^^^ | -1.49  (-2.26, -0.73) | 2.11  (-0.73, 4.94) | -^&^ | -2.28  (-2.61, -1.95) | 2.65  (1.12, 4.17) | -2.67  (-4.18, -1.16) |
|  | Median (IQR) of estimates^^*^ | -1.52 (0.77) | 2.17 (3.28) | -^&^ | -2.29 (0.29) | 2.66 (1.30) | -2.67 (1.18) |
|  | Mean (MSE) of estimates^^*^ | -1.14 (5.38) | 2.25 (9.05) | -^&^ | -2.27 (0.02) | 2.63 (0.44) | -2.68 (0.39) |
| TZH | True parameters  (95% CI)^^^ | -1.95  (-2.55, -1.35) | 2.00  (-0.68, 4.67) | -3.18  (-5.52, -0.84) | -2.18  (-2.53, -1.83) | 7.80  (6.24, 9.37) | -6.07  (-7.57, -4.58) |
|  | Median (IQR) of estimates^^*^ | -1.94 (0.61) | 2.01 (2.39) | -3.15 (1.93) | -2.21 (0.33) | 7.94 (1.51) | -6.16 (1.43) |
|  | Mean (MSE) of estimates^^*^ | -1.93 (0.05) | 1.95 (0.77) | -3.12 (0.60) | -2.18 (0.03) | 7.91 (0.53) | -6.21 (0.53) |

^&^Given preliminary analysis indicated an identifiability issue existed in the model when parameter c was kept in the model for the data at this site, this parameter was excluded for formal analysis.

^^^Parameters estimated from the observed data were described as true parameters in the simulation study. With the true parameters, we simulated 100 epidemics of *Campylobacter* spp. infection for each site. We fitted the Markov model to these 100 epidemics to estimate the parameters for each epidemic, described as estimated parameters, and reported their medians and means.

^*^SAV – Venda, South Africa; TZH – Haydom, Tanzania; IQR – interquartile range; MSE – mean squared error.

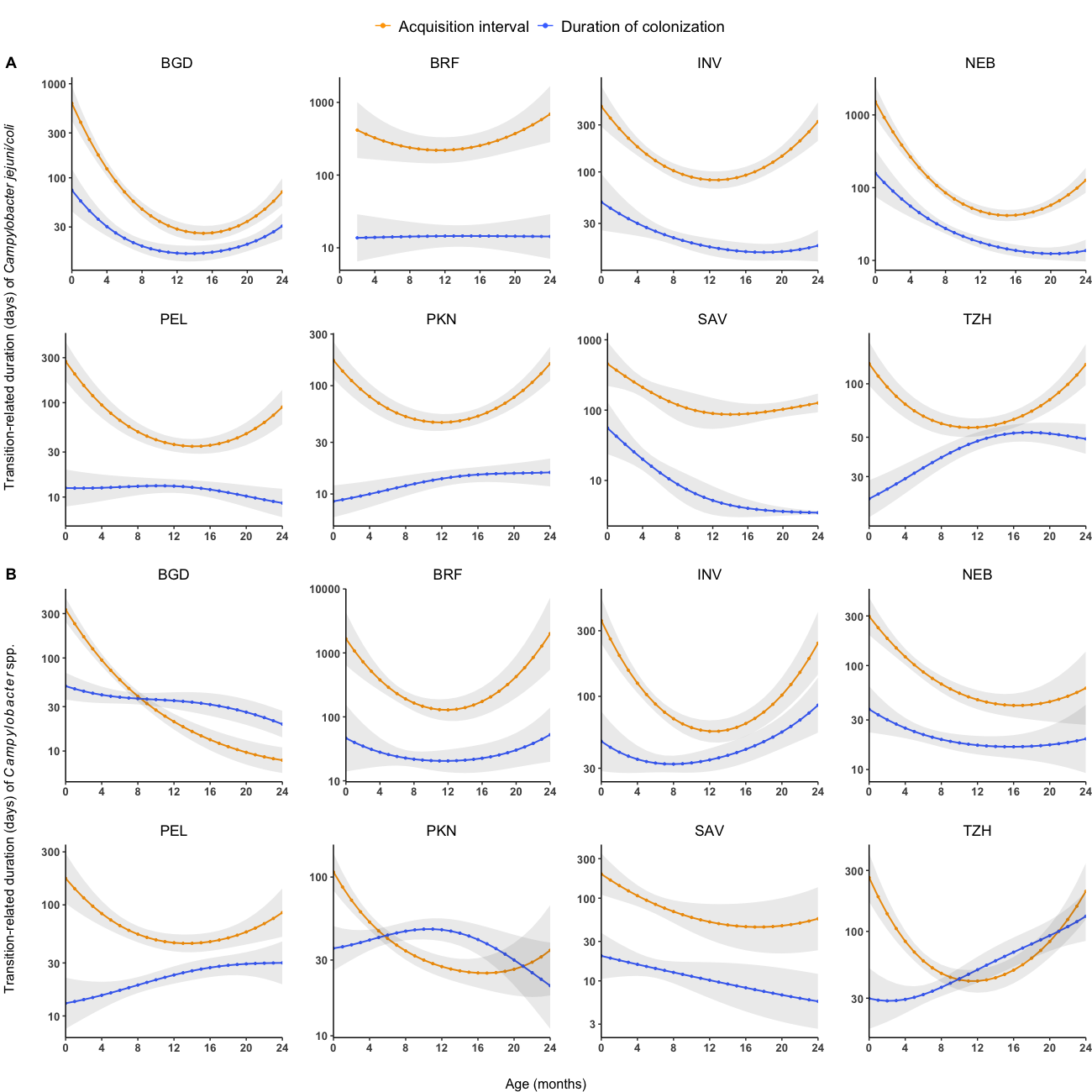

**Supplementary figure 1** Model-estimated site-specific age-dependent mean time to acquisition ($\frac{\boldsymbol{1}}{\boldsymbol{p}_{\boldsymbol{t}}}$, orange) and mean time to clearance ($\frac{\boldsymbol{1}}{\boldsymbol{q}_{\boldsymbol{t}}}$, blue) for *Campylobacter* *jejuni/coli* (A) and *Campylobacter* spp. (B). Grey shades are 95% asymptotic confidence bands. Sites included: Dhaka, Bangladesh (BGD); Vallore, India (INV); Bhaktapur, Nepal (NEB); Naushero Feroze, Pakistan (PKN); Venda, South Africa (SAV); Haydom, Tanzania (TZH); Fortaleza, Brazil (BRF); Loreto, Peru (PEL).

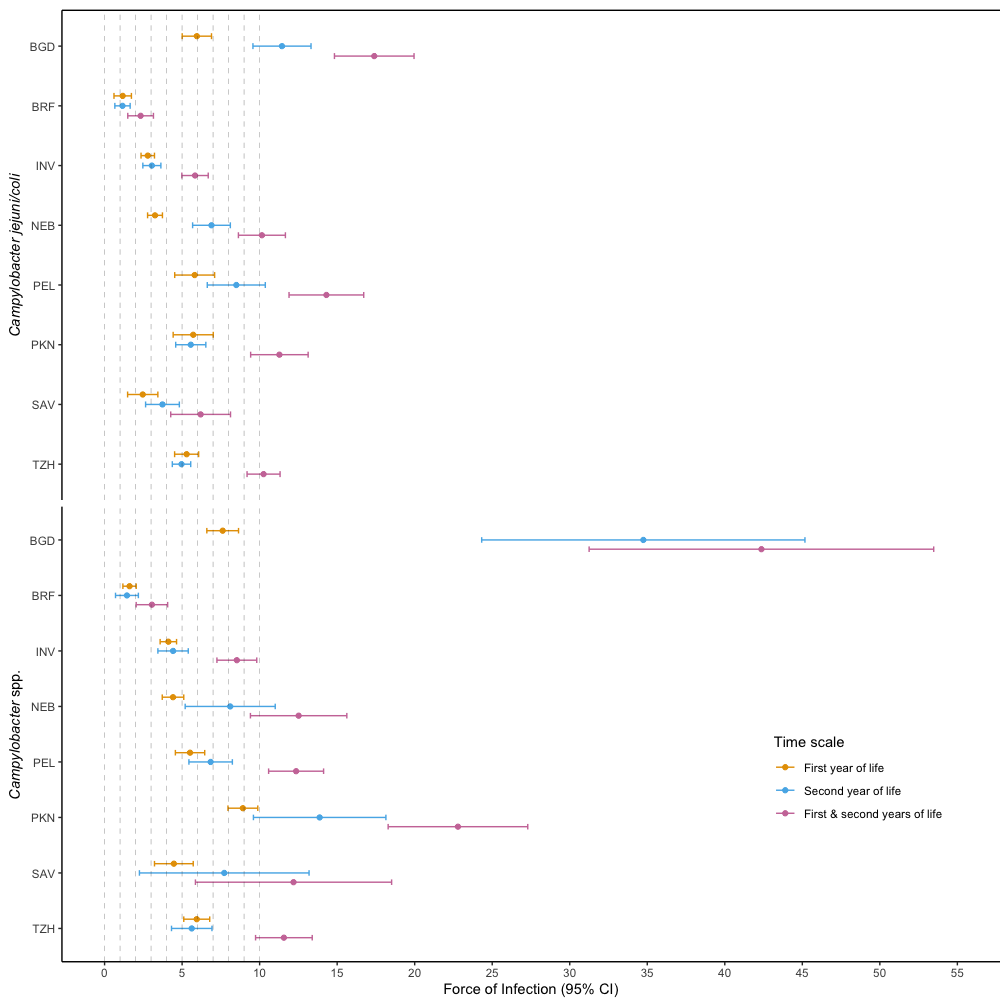

**Supplementary figure 2** Site-specific forces of infection of *Campylobacter* *jejuni/coli* and *Campylobacter* spp. Sites included: Dhaka, Bangladesh (BGD); Vallore, India (INV); Bhaktapur, Nepal (NEB); Naushero Feroze, Pakistan (PKN); Venda, South Africa (SAV); Haydom, Tanzania (TZH); Fortaleza, Brazil (BRF); Loreto, Peru (PEL).

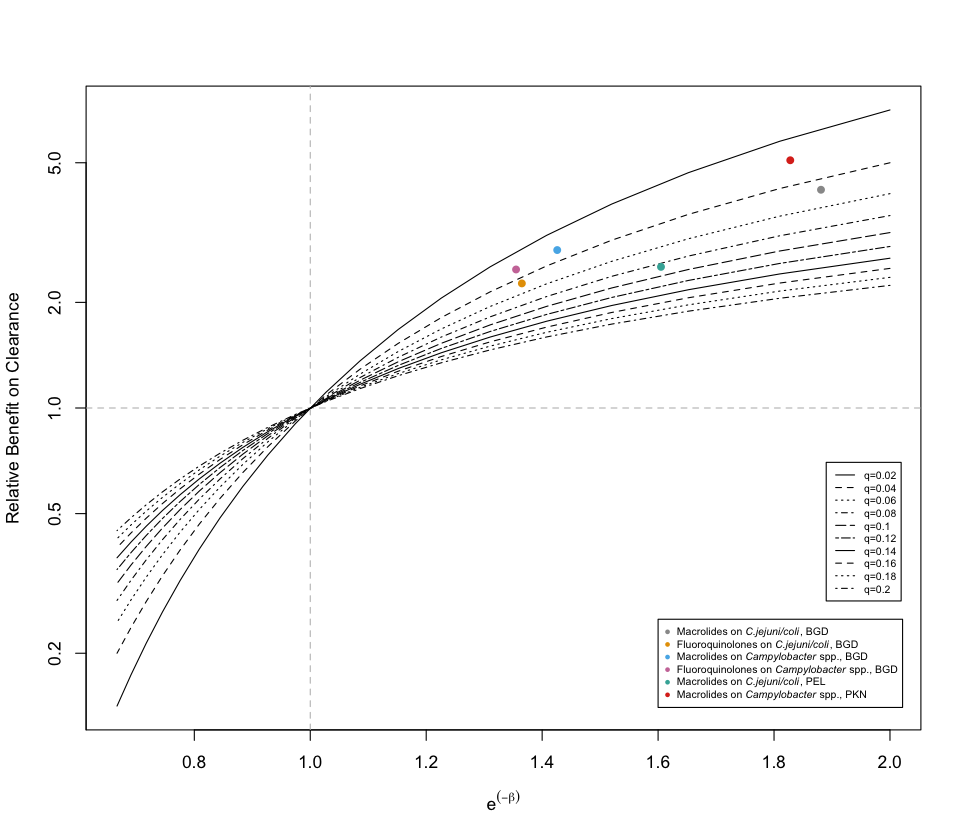

**Supplementary figure 3** Relationship between relative benefit $\frac{q_{t}\left( x\left( t \right)=1 \right)}{q_{t}\left( x\left( t \right)=0 \right)}$ and the acceleration factor for clearance (AFC), exp(-$\beta$), at various levels of baseline clearance probability$q_{t}\left( x\left( t \right)=0 \right)$. We use AFC for the evaluation of antibiotic effectiveness on clearance of *Campylobacter*. The relative benefits of the significant AFCs, based on the mean of daily clearance probabilities of the 24-month follow-up by *Campylobacter* group and site, are marked by colored point. Sites included: Dhaka, Bangladesh (BGD); Naushero Feroze, Pakistan (PKN); Loreto, Peru (PEL).

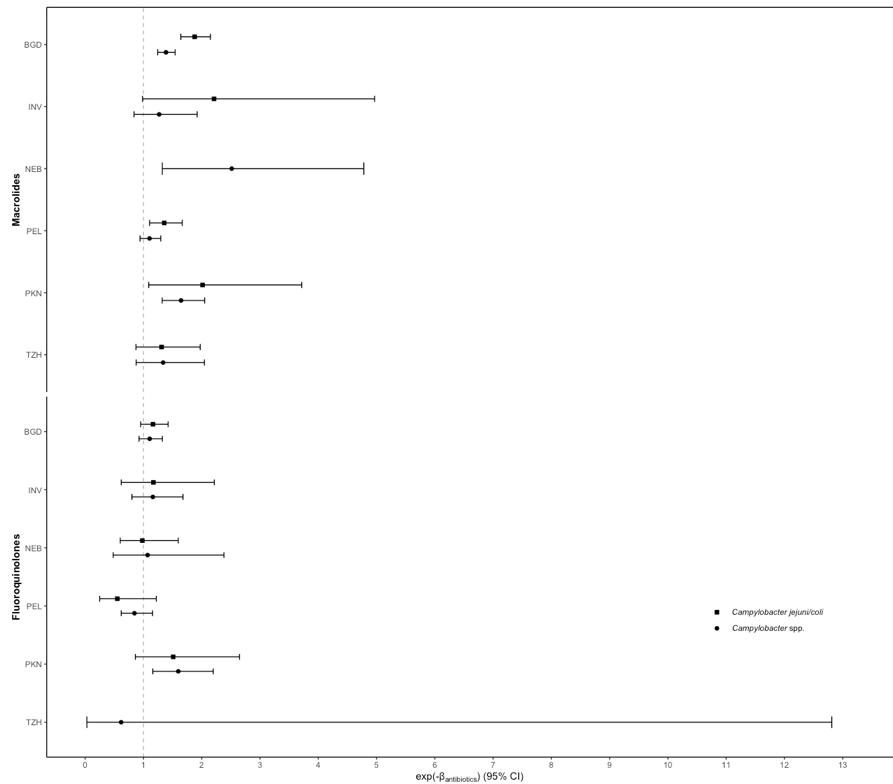

**Supplementary figure 4** Site-specific effects of prior fifteen-day usage of macrolides and fluoroquinolones on the clearances of *Campylobacter jejuni/coli* and *Campylobacter* spp. Point estimates and 95% confidence intervals of the acceleration factor for clearance (AFC) were shown, which is defined as the ratio of the log probability of clearance with antibiotic use to the log probability of clearance without. Sites included: Dhaka, Bangladesh (BGD); Vallore, India (INV); Bhaktapur, Nepal (NEB); Naushero Feroze, Pakistan (PKN); Venda, South Africa (SAV); Haydom, Tanzania (TZH); Fortaleza, Brazil (BRF); Loreto, Peru (PEL).
